# Familial risk of psychosis in obsessive-compulsive disorder: Impact on clinical characteristics, comorbidity and treatment response

**DOI:** 10.1101/2022.04.17.22273465

**Authors:** Srinivas Balachander, Navya Spurthi Thatikonda, Anand Jose Kannampuzha, Mahashweta Bhattacharya, Sweta Sheth, Vinutha Ramesh, Alen Chandy Alexander, Muthukumaran Moorthy, Mino Susan Joseph, Sowmya Selvaraj, Dhruva Ithal, Vanteemar S Sreeraj, John P John, Ganesan Venkatasubramanian, Biju Viswanath, YC Janardhan Reddy, Sanjeev Jain, ADBS consortium

## Abstract

**Background:** Family studies in obsessive-compulsive disorder (OCD) indicate higher rates of psychosis among their first-degree relatives (FDRs). However, the etiological and clinical relationships between the two disorders remain unclear. We compared the clinical characteristics & pharmacological treatment response in patients diagnosed with OCD with a family history of psychosis (OCD-FHP), with a family history of OCD (OCD-FHO) and those with sporadic OCD (OCD-S).

**Methods:** A total of 226 patients who met DSM-IV criteria for OCD (OCD-FHP=59, OCD-FHO=112, OCD-S=55) were included for analysis. All patients were evaluated using the Mini International Neuropsychiatric Interview (MINI 6.0.0), Yale-Brown Obsessive-Compulsive Scale (YBOCS), and the Family Interview for Genetic Studies (FIGS). Treatment response was characterized over naturalistic follow-up.

**Results:** The three groups did not differ across any demographic or clinical variables other than treatment response. Patients in the OCD-FHP group were found to have received a greater number of trials with serotonin reuptake inhibitors (SRI) [F(2,223)=7.99, p<0.001], were more likely to have failed ≥ 2 trials of SRIs (χ^2^=8.45, p=0.014), and less likely to have attained remission (χ^2^=6.57, p=0.037)

**Conclusions:** We observed that having a relative with psychosis may predispose to treatment resistance in OCD. Further research on the influence of genetic liability to psychosis on treatment response in OCD may offer novel translational leads.

## 1. Introduction

Obsessive-Compulsive Disorder (OCD) is a complex neuropsychiatric illness with a prevalence of 1-3% (Fontenelle et al., 2006). OCD is heterogeneous in its clinical presentation and manifests a wide range of clinical symptoms, course, and treatment responses. Several factors are understood to influence the presentation of OCD, such as age at onset (Kichuk et al., 2013; Narayanaswamy et al., 2012), comorbidities such as tics or mood disorders (Conelea et al., 2014; Jaisoorya et al., 2008; Viswanath et al., 2012) and family history (Arumugham et al., 2014; Hanna et al., 2005). It is suggested that this heterogeneous mix of clinical phenotypes reflects a wide range of genetic vulnerabilities (Mahjani et al., 2021).

The relationship between psychosis and OCD has piqued interest for several years (Roth 1960) due to many reasons. There is a higher-than-expected rate of OCD in samples of schizophrenia (12-14%) and a much higher occurrence of subsyndromal OC symptoms (30%) (Swets et al., 2014). Conversely, several registry-based longitudinal studies in OCD have found a higher risk of schizophrenia & other psychotic disorders in those who receive an initial diagnosis of OCD (Cheng et al., 2019; Meier et al., 2014; Van Dael et al., 2011). Also, family studies looking at OCD & schizophrenia have found a higher prevalence of either disorder among first-degree relatives (Cederlöf et al., 2015; Poyurovsky et al., 2005).

The clinical presentation of OCD co-morbid with schizophrenia & other psychotic disorders has also been studied (Devi et al., 2015; Faragian et al., 2009; Rajkumar et al., 2008). Most of these studies have only looked at how the presence of OCD/OCS may impact the clinical presentation and course of schizophrenia, but not that of OCD. One study did compare the clinical characteristics of OCD, with and without comorbid schizophrenia (Ozdemir et al., 2003), which found that both groups did not differ in terms of phenomenology or in terms of severity & insight into the OCD symptoms. However, the study was limited by its small sample size (20 vs 45). Many studies in OCD samples observe that the presence of schizotypy or comorbid schizotypal disorder is often associated with a different symptom profile (Brakoulias et al., 2014), lower levels of insight (Poyurovsky et al., 2008) and poorer treatment response (Perris et al., 2019).

As dysfunction in the cortico-striato-thalamic circuitry is postulated to be a key mechanism in both disorders, there is also substantial overlap in the endophenotypes identified between them. For example, many studies in OCD have reported working memory deficits, which is considered a key endophenotype in schizophrenia (Abramovitch et al., 2015; Gur et al., 2015). Aberrations in certain neurophysiological measures such as evoked potentials (P300, mismatch negativity, etc.) are common to both disorders. Several genetic variants that contribute to risk are shared between OCD and schizophrenia (Qin et al., 2016). More recently, cross-disorder genetic studies seem to suggest that genetic variants from genome-wide association studies (GWAS) are also shared across the two disorders (Brainstorm Consortium et al., 2018), with cross-heritability and genetic correlation estimate of around 0.34 (Smoller et al., 2019). This suggests that there may be common pathophysiological mechanisms underlying both disorders.

A familial liability for psychosis, could thus influence clinical manifestations such as symptom profile, comorbidities, course & outcomes in OCD. A recent registry-based study on eating disorders found that a family history of schizophrenia was associated with distinct patterns of comorbidity, outcomes and illness burden (Zhang R et al., 2021). Similar studies in mood disorders (Coryell et al., 1985; Maj et al., 1991; Pavuluri et al., 2004; Vandeleur et al., 2014), have been of immense heuristic value in improving our understanding of the psychosis continuum - specifically between affective psychoses, schizoaffective disorders and schizophrenia.

We thus compared a sample of subjects with familial risk of psychosis versus those with familial risk of OCD and a sample of sporadic OCD. We investigated whether the three groups might differ based on clinical characteristics such as symptom dimensions, insight, comorbidity, age at onset, course & treatment outcomes.

## 2. Materials & Methods

### 2.1 Subjects & Recruitment

Subjects belonging to the familial OCD (OCD-FHO) and the OCD with a family history of psychosis (OCD-FHP) groups were recruited as part of the Accelerator Program for Discovery in Brain disorders using Stem cells (ADBS) (Viswanath et al., 2018), an ongoing longitudinal study looking at multiplex affected families. We approached all treatment-seeking individuals with a primary diagnosis of OCD, identified during the routine clinical examination to have a family history of any major psychiatric illness in any first-degree relative for participation. The diagnosis of DSM-IV-TR OCD was established using the Mini International Neuropsychiatric Interview version 6.0.0 (Sheehan et al., 1998), and family history was ascertained using the Family Interview for Genetic Studies (FIGS) (Maxwell, 1996). As the original FIGS did not have a module for OCD, we modified questions from the OCD section of the MINI, re-framed them to inquire about a family history of OCD and included this as part of the FIGS interview. We interviewed at least three members from each family to ascertain the family history in both first- and second-degree relatives. If an affected family member could not be interviewed directly, a diagnosis was inferred only if the information from all the three available family members was corroborated. Further details regarding the nature of the ADBS sample and other inclusion/exclusion criteria have been reported elsewhere (Sreeraj et al., 2020). For the current study, patients in the OCD-FHP group were those who had a primary diagnosis of OCD, and at least one first-degree relative who was diagnosed with either schizophrenia or any other primary psychotic disorder. Those in the OCD-FHO group, had a first-degree relative affected with OCD, and no first or second-degree relative affected with schizophrenia or other psychoses.

“Sporadic” OCD (OCD-S) was defined as those with OCD who did not have any family history of a major psychiatric illness in their first or second-degree relatives. The OCD-S sample was recruited around the same time frame as the other two groups as part of an independent, parallel study aimed at investigating the neurocognitive endophenotypes between the familial and sporadic groups (Bhattacharya et al., 2021). Written informed consent was obtained from all participants, and the Institute Ethics Committee approved the study.

### 2.2 Assessments

Symptom profile & severity were assessed using the Yale-Brown Obsessive-Compulsive Scale (YBOCS) (Goodman et al., 1989b, 1989a). Insight was assessed using item-11 of the YBOCS. Psychiatric comorbidities were evaluated using the MINI 6.0.0 plus (Sheehan et al., 1998). Their treatment-related details, specifically symptom response to pharmacological treatments, were collected from their medical records over naturalistic follow-up. Treatment response to serotonin reuptake inhibitors (SRIs) was assessed using international consensus criteria (Mataix-Cols et al., 2019), by which an SRI trial was considered failed if there was <25% reduction in the YBOCS total score and a CGI-I of >2 even after 8-12 weeks at therapeutic doses. Remission was defined if the YBOCS score was <12, with a CGI-S or 1 or 2. All assessments were done by clinicians with expertise in assessing and treating OCD and regularly trained in the use of the YBOCS.

### 2.3 Statistical Analysis

Statistical comparisons between the three groups OCD with familial risk of psychosis (OCD-FHP), familial OCD (OCD-FHO) and sporadic OCD (OCD-S) was carried out using analysis of variance (ANOVA) for continuous measures and chi-square tests for dichotomous measures. To adjust for multiple comparisons, we used Bonferroni correction both for the omnibus tests (by setting the p-value threshold as 0.05 divided by the number of tests carried out within that domain), and the post-hoc pairwise tests. All analyses were done in R version 4.1.1 (R Core Team, 2021), using the base packages.

## 3. Results

The subjects for the familial groups (OCD-FHP & OCD-FHO) were taken from the ADBS study database, which had 224 probands with a diagnosis of OCD. Of these, 34 were excluded as they had a primary/comorbid diagnosis of schizophrenia, and an additional 19 were excluded as they had comorbid bipolar disorder. Thus, we compared the OCD-FHP group (n=59), OCD-FHO (n=112) with the OCD-S sample (n=55). Table 1 shows the results of these comparisons with respect to the demographic, symptom profile, severity, and comorbidity. No significant differences were seen with any of the demographic or clinical characteristics with respect to symptoms & comorbidity profile. Table 2 shows the comparisons of the baseline & current illness severity, and the treatment response between the three groups. The OCD-FHP group was found to have a greater mean number of SRI trials (F=7.992, p<0.001) compared to both OCD-FHO and the OCD-S group (post-hoc pairwise tests showed OCD-FHP>OCD-FHO=OCD-S). Similarly, SRI-resistance (i.e., failed ≥ 2 adequate SRI trials) was seen in higher proportion in the OCD-FHP group (36%), as compared to the OCD-FHO group (18%) and OCD-S group (16%) (χ^2^ =8.455; p=0.014). Also, lesser rates of remission were found in the OCD-FHP group, as compared to the other two. Though only the first comparison (number of SRI trials) was found to survive multiple comparison correction, the other results also point to the general direction of greater treatment resistance or attenuated response to SRIs in the OCD-FHP group.

**Table 1.** Differences in demographic, clinical characteristics, comorbidity profile & treatment response comparing the three groups.

| Variable | OCD with family history of psychosis (n=59) | Familial OCD, without family history of psychosis (n=112) | Sporadic OCD (n=55) | F (2,223) or $\chi^2$ | P-value |
| --- | --- | --- | --- | --- | --- |
| **Socio-demographic** |  |  |  |  |  |
| Age | 35.76 (9.82) | 36.13 (7.43) | 32.75 (11.22) | 2.663 | 0.072 |
| Sex (male) | 36 (61%) | 58 (52%) | 29 (53%) | 1.412 | 0.494 |
| Age at Onset | 19.59 (9.72) | 20.81 (8.58) | 22.53 (9.92) | 1.155 | 0.317 |
| Duration of illness (in years) | 13.46 (7.84) | 12.36 (7.29) | 10.24 (7.97) | 1.73 | 0.18 |
| Years of Education | 13.2 (3.61) | 12.1 (4.62) | 13.8 (2.7) | 2.417 | 0.092 |
| Married | 20 (34%) | 58 (52%) | 29 (53%) | 5.804 | 0.055 |
| **YBOCS Checklist Items (Lifetime)** |  |  |  |  |  |
| **Obsessions** |  |  |  |  |  |
| Contamination | 29 (49%) | 72 (64%) | 33 (60%) | 3.682 | 0.159 |
| Somatic | 7 (12%) | 11 (10%) | 4 (7%) | 0.685 | 0.767<sup>a</sup> |
| Aggressive | 17 (29%) | 33 (29%) | 15 (27%) | 0.087 | 0.957 |
| Sexual | 14 (24%) | 27 (24%) | 9 (16%) | 1.403 | 0.496 |
| Religious | 15 (25%) | 29 (26%) | 9 (16%) | 2.039 | 0.361 |
| Pathological Doubts | 23 (39%) | 47 (42%) | 19 (35%) | 0.856 | 0.652 |
| Hoarding | 4 (7%) | 11 (10%) | 1 (2%) | 3.602 | 0.175<sup>a</sup> |
| Need for Symmetry | 15 (25%) | 29 (26%) | 13 (24%) | 0.101 | 0.951 |
| Miscellaneous Obsessions | 16 (27%) | 33 (29%) | 11 (20%) | 1.708 | 0.426 |
| **Compulsions** |  |  |  |  |  |
| Washing | 29 (49%) | 69 (62%) | 32 (58%) | 2.466 | 0.291 |
| Checking | 26 (44%) | 48 (43%) | 20 (36%) | 0.842 | 0.657 |
| Ordering/Arranging | 11 (19%) | 24 (21%) | 10 (18%) | 0.324 | 0.850 |
| Repeating | 14 (24%) | 28 (25%) | 14 (25%) | 0.051 | 0.974 |
| Counting | 9 (15%) | 15 (13%) | 4 (7%) | 1.876 | 0.391<sup>a</sup> |
| Collecting | 4 (7%) | 6 (5%) | 0 (0%) | 3.550 | 0.152<sup>a</sup> |
| Miscellaneous Compulsions | 32 (54%) | 53 (47%) | 20 (36%) | 3.722 | 0.156 |
| **Comorbidity (Lifetime)** |  |  |  |  |  |
| Major Depressive Disorder | 9 (15%) | 24 (20%) | 18 (33%) | 5.919 | 0.077 |
| Dysthymia | 2 (3%) | 7 (6%) | 6 (11%) | 2.651 | 0.298<sup>a</sup> |
| Panic Disorder | 2 (3%) | 4 (4%) | 5 (9%) | 5.059 | 0.111<sup>a</sup> |
| Agoraphobia | 0 (0%) | 1 (1%) | 1 (2%) | - | - |
| **Social Anxiety Disorder** | 2 (3%) | 10 (9%) | 6 (11%) | 5.122 | 0.112<sup>a</sup> |
| **Alcohol Use Disorder** | 0 (0%) | 0 (0%) | 1 (2%) | - | - |
| **Other substance use disorders** | 1 (2%) | 1 (1%) | 0 (0%) | - | - |
| **Generalized Anxiety Disorder** | 0 (0%) | 3 (3%) | 6 (11%) | - | - |
| **Tic Disorder** | 2 (4%) | 0 (0%) | 3 (5%) | - | - |
| **Body Dysmorphic Disorder** | 1 (2%) | 0 (0%) | 0 (0%) | - | - |
| **Attention-Deficit Hyperactivity Disorder** | 1 (2%) | 1 (1%) | 0 (0%) | - | - |

*a- Fisher Exact test; OCD – Obsessive-Compulsive Disorder*

**Table 2.** Comparison of severity of OCD and treatment response between the three groups.

| Variable | OCD with family history of psychosis (n=59) | Familial OCD, without family history of psychosis (n=112) | Sporadic OCD (n=55) | F(2,223) or Chi square | P-value |
| --- | --- | --- | --- | --- | --- |
| Duration of follow-up in months (median & range) | 80 (9 - 126) | 68 (12 - 147) | 72 (14 - 205) | - | - |
| Episodic OCD Course | 1 (2%) | 3 (3%) | 4 (7%) | 3.076 | 0.322<sup>a</sup> |
| **Baseline Severity** |  |  |  |  |  |
| YBOCS Obsessions Score | 12.97 (4.09) | 12.32 (4.04) | 11.92 (4.26) | 0.749 | 0.474 |
| YBOCS Compulsions Score | 11.46 (5.32) | 11.97 (4.62) | 10.61 (5.49) | 0.445 | 0.641 |
| YBOCS Total Score | 24.65 (8.19) | 24.12 (8.11) | 22.59 (8.77) | 0.791 | 0.454 |
| Poor Insight | 12 (20%) | 12 (11%) | 6 (10%) | 3.884 | 0.143 |
| CGI-S | 4.72 (1.04) | 4.49 (1.02) | 4.39 (1.09) | 1.751 | 0.176 |
| **Current Severity** |  |  |  |  |  |
| YBOCS Obsessions Score | 8.73 (3.85) | 10.49 (4.39) | 9.25 (4.57) | 2.151 | 0.119 |
| YBOCS Compulsions Score | 7.92 (3.21) | 9.52 (5.28) | 7.96 (4.41) | 1.644 | 0.195 |
| YBOCS Total Score | 16.21 (7.89) | 18.91 (9.41) | 17.23 (8.89) | 0.848 | 0.430 |
| Poor Insight | 6 (10%) | 4 (4%) | 2 (4%) | 3.751 | 0.187<sup>a</sup> |
| CGI-S | 3.70 (1.29) | 3.73 (1.28) | 3.17 (1.34) | 1.051 | 0.351 |
| Responders | 30 (51%) | 67 (60%) | 35 (64%) | 5.773 | 0.217 |
| Partial Responders | 22 (37%) | 25 (22%) | 14 (25%) |  |  |
| **Remitters** | **19 (32%)** | **55 (49%)** | **30 (55%)** | **6.573** | **0.037** |
| **Current Medication Trials** |  |  |  |  |  |
| Any SRI | 59 (100%) | 109 (97%) | 53 (96%) | 1.963 | 0.374 |
| Fluoxetine | 18 (31%) | 25 (22%) | 15 (27%) | 0.382 | 0.744 |
| Escitalopram | 14 (24%) | 29 (26%) | 12 (22%) |  |  |
| Sertraline | 5 (8%) | 16 (14%) | 11 (20%) |  |  |
| Paroxetine | 0 (0%) | 1 (1%) | 3 (5%) |  |  |
| Fluvoxamine | 4 (7%) | 13 (12%) | 4 (7%) |  |  |
| Clomipramine | 10 (17%) | 15 (13%) | 5 (9%) |  |  |
| Antipsychotic augmentation | 19 (32%) | 20 (18%) | 11 (20%) | 5.209 | 0.074 |
| **Number of SRI trials** | **1.92 (0.88)** | **1.35 (0.91)** | **1.17 (0.61)** | **7.992** | **<0.001** |
| **Failed ≥2 SRI trials** | **21 (36%)** | **20 (18%)** | **9 (16%)** | **8.455** | **0.014** |

*a- Fisher Exact test; OCD – Obsessive-Compulsive Disorder; YBOCS – Yale-Brown Obsessive-Compulsive Scale; CGI-S – Clinical Global Impression – Severity; SRI – Serotonin Reuptake Inhibitor*

## 4. Discussion

The main aim of this study was to understand whether a familial risk of psychosis might influence the clinical presentation, severity, comorbidity or pharmacological treatment response in OCD. Based on existing family studies & GWAS, both disorders are likely to have shared genetic risk factors. Our study’s main finding is the higher rates of pharmacological (SRI) resistance in the group with a family history of psychosis. Also, the clinical presentation, symptomatology, severity or comorbidity rates were not different between the three groups.

Existing research seems to indicate that the clinical presentation of OCD in comorbid schizophrenia appears to be similar to OCD without comorbid schizophrenia (Ozdemir et al., 2003). Still, no study has yet evaluated treatment outcomes. Our study’s findings appear to reflect these observations regarding the presentation of OCD in those with a familial risk of schizophrenia. Our observation concerning the attenuated treatment response to SRI treatment in the OCD group with familial risk of psychosis has several implications, both clinical and neurobiological.

Treatment response in OCD is also known to be very heterogeneous, with around 40-60% of OCD patients being resistant to treatment with SRIs. The reasons for SRI resistance are largely unknown, and there are few consistent predictors for it (Hazari et al., 2018). One consistent predictor of poor outcome in OCD is schizotypal personality, which is also associated with a family history of schizophrenia. Other features of OCD with comorbid schizotypal personality include greater illness severity, poorer insight, lower resistance & control to OCD symptoms. In our sample, we assessed for symptoms of psychosis using the structured MINI, and we excluded patients with a comorbid psychotic disorder. This could be a reason why we did not find differences in symptomatology or severity in the samples but only in treatment response. Further, as there were also no other confounding differences between the groups (such as differences in gender distribution, duration of illness, other comorbidities, etc) this may indeed be a robust finding indicating the influence of familial risk on treatment response.

Both OCD and schizophrenia have dysfunction in the serotonergic and dopaminergic systems at the neurotransmitter level. While the primary pharmacological agents used for OCD are SRIs, D2-blocking antipsychotics are a first-line augmentation strategy. We observed a non-significant trend towards higher rates of antipsychotic use in the OCD-FHP group; we did not have sufficient information to comment on the response to antipsychotics. The genetic vulnerability for psychoses might influence neurotransmitter systems, to adversely impact response to SRIs. Also, a recent GWAS study in major depressive disorder found that the polygenic risk score (PRS) of schizophrenia predicted a poorer response to antidepressant use (Pain et al., 2020). Similar approaches in OCD (i.e., using PRS for psychosis/schizophrenia in OCD samples) to predict treatment response may help identify more specific markers for poor SRI response.

The main strengths of our study are its unique sample profile, well-characterized clinical phenotype, treatment response and family profile. However, there are several limitations: we did not have an assessment of schizotypal traits. Our treatment response was ascertained through a naturalistic follow-up, which is inherently fraught with several confounding factors. Nonetheless, adherence and treatment response are verified by experienced psychiatrists, with corroborative reliable clinical information from caregivers who are most often first-degree relatives. As the study was exploratory, it is unclear if the sample size was sufficient to detect differences between the groups.

## 5. Conclusions

In summary, the familial risk of psychosis in OCD appears only to affect SRI treatment response and not other aspects in OCD. The clinical implication of this finding might be the predictive value of familial psychosis risk in prognosticating SRI response. Including a detailed family history as part of clinical examination could potentially inform treatment decisions. Whether OCD patients with familial risk of psychosis might benefit better from using other pharmacological agents (such as anti-dopaminergic or glutamatergic agents), merits further study. Future research on pharmacological treatment response in OCD could take this into consideration, and genetic studies need to account for how familial aggregation of these syndromes may influence one another.

## Data Availability

All data produced in the present study are available upon reasonable request to the corresponding

## Acknowledgements

Nil

## References

Abramovitch, A., Mittelman, A., Tankersley, A.P., Abramowitz, J.S., Schweiger, A., 2015. Neuropsychological investigations in obsessive-compulsive disorder: A systematic review of methodological challenges. Psychiatry Res 228, 112–120. https://doi.org/10.1016/j.psychres.2015.04.025

Arumugham, S.S., Cherian, A.V., Baruah, U., Viswanath, B., Narayanaswamy, J.C., Math, S.B., Reddy, Y.C.J., 2014. Comparison of clinical characteristics of familial and sporadic obsessive-compulsive disorder. Compr Psychiatry 55, 1520–1525. https://doi.org/10.1016/j.comppsych.2014.07.006

Bhattacharya, M., Balachander, S., Viswanath, B., Reddy, Y.C.J., 2021. Comparison of neurocognitive performance in familial versus sporadic obsessive-compulsive disorder. Journal of Obsessive-Compulsive and Related Disorders 30, 100666. https://doi.org/10.1016/j.jocrd.2021.100666

Brainstorm Consortium, Anttila, V., Bulik-Sullivan, B., Finucane, H.K., Walters, R.K., Bras, J., Duncan, L., Escott-Price, V., Falcone, G.J., Gormley, P., Malik, R., Patsopoulos, N.A., Ripke, S., Wei, Z., Yu, D., Lee, P.H., Turley, P., Grenier-Boley, B., Chouraki, V., Kamatani, Y., …, Murray, R., 2018. Analysis of shared heritability in common disorders of the brain. Science 360, eaap8757. https://doi.org/10.1126/science.aap8757

Brakoulias, V., Starcevic, V., Berle, D., Milicevic, D., Hannan, A., Viswasam, K., Mann, K., 2014. The clinical characteristics of obsessive compulsive disorder associated with high levels of schizotypy. Aust N Z J Psychiatry 48, 852–860. https://doi.org/10.1177/0004867414531831

Cederlöf, M., Lichtenstein, P., Larsson, H., Boman, M., Rück, C., Landén, M., Mataix-Cols, D., 2015. Obsessive-Compulsive Disorder, Psychosis, and Bipolarity: A Longitudinal Cohort and Multigenerational Family Study. Schizophr Bull 41, 1076–1083. https://doi.org/10.1093/schbul/sbu169

Cheng, Y.-F., Chen, V.C.-H., Yang, Y.-H., Chen, K.-J., Lee, Y.-C., Lu, M.-L., 2019. Risk of schizophrenia among people with obsessive-compulsive disorder: A nationwide population-based cohort study. Schizophrenia Research 209, 58–63. https://doi.org/10.1016/j.schres.2019.05.024

Conelea, C.A., Walther, M.R., Freeman, J.B., Garcia, A.M., Sapyta, J., Khanna, M., Franklin, M., 2014. Tic-related obsessive-compulsive disorder (OCD): phenomenology and treatment outcome in the Pediatric OCD Treatment Study II. J Am Acad Child Adolesc Psychiatry 53, 1308–1316. https://doi.org/10.1016/j.jaac.2014.09.014

Coryell, W., Endicott, J., Keller, M., Andreasen, N.C., 1985. Phenomenology and family history in DSM-III psychotic depression. J Affect Disord 9, 13–18. https://doi.org/10.1016/0165-0327(85)90004-7

Devi, S., Rao, N.P., Badamath, S., Chandrashekhar, C.R., Janardhan Reddy, Y.C., 2015. Prevalence and clinical correlates of obsessive-compulsive disorder in schizophrenia. Compr Psychiatry 56, 141–148. https://doi.org/10.1016/j.comppsych.2014.09.015

Faragian, S., Pashinian, A., Fuchs, C., Poyurovsky, M., 2009. Obsessive-compulsive symptom dimensions in schizophrenia patients with comorbid obsessive-compulsive disorder. Prog Neuropsychopharmacol Biol Psychiatry 33, 1009–1012. https://doi.org/10.1016/j.pnpbp.2009.05.008

Fontenelle, L.F., Mendlowicz, M.V., Versiani, M., 2006. The descriptive epidemiology of obsessive-compulsive disorder. Prog. Neuropsychopharmacol. Biol. Psychiatry 30, 327–337. https://doi.org/10.1016/j.pnpbp.2005.11.001

Goodman, W.K., Price, L.H., Rasmussen, S.A., Mazure, C., Fleischmann, R.L., Hill, C.L., Heninger, G.R., Charney, D.S., 1989. The Yale-Brown Obsessive Compulsive Scale. I. Development, use, and reliability. Arch. Gen. Psychiatry 46, 1006–1011.

Goodman, W.K., Price, L.H., Rasmussen, S.A., Mazure, C., Delgado, P., Heninger, G.R., Charney, D.S., 1989. The Yale-Brown Obsessive Compulsive Scale. II. Validity. Arch. Gen. Psychiatry 46, 1012–1016.

Gur, R.C., Braff, D.L., Calkins, M.E., Dobie, D.J., Freedman, R., Green, M.F., Greenwood, T.A., Lazzeroni, L.C., Light, G.A., Nuechterlein, K.H., Olincy, A., Radant, A.D., Seidman, L.J., Siever, L.J., Silverman, J.M., Sprock, J., Stone, W.S., Sugar, C.A., Swerdlow, N.R., Tsuang, D.W., Tsuang, M.T., Turetsky, B.I., Gur, R.E., 2015. Neurocognitive performance in family-based and case-control studies of schizophrenia. Schizophr. Res. 163, 17–23. https://doi.org/10.1016/j.schres.2014.10.049

Hanna, G.L., Himle, J.A., Curtis, G.C., Gillespie, B.W., 2005. A family study of obsessive-compulsive disorder with pediatric probands. Am. J. Med. Genet. B Neuropsychiatr. Genet. 134B, 13–19. https://doi.org/10.1002/ajmg.b.30138

Jaisoorya, T.S., Reddy, Y.C.J., Srinath, S., Thennarasu, K., 2008. Obsessive-compulsive disorder with and without tic disorder: a comparative study from India. CNS Spectr 13, 705–711. https://doi.org/10.1017/s1092852900013791

Kichuk, S.A., Torres, A.R., Fontenelle, L.F., Rosário, M.C., Shavitt, R.G., Miguel, E.C., Pittenger, C., Bloch, M.H., 2013. Symptom dimensions are associated with age of onset and clinical course of obsessive-compulsive disorder. Prog Neuropsychopharmacol Biol Psychiatry 44, 233–239. https://doi.org/10.1016/j.pnpbp.2013.02.003

Mahjani, B., Klei, L., Mattheisen, M., Halvorsen, M.W., Reichenberg, A., Roeder, K., Pedersen, N.L., Boberg, J., de Schipper, E., Bulik, C.M., Landén, M., Fundín, B., Mataix-Cols, D., Sandin, S., Hultman, C.M., Crowley, J.J., Buxbaum, J.D., Rück, C., Devlin, B., Grice, D.E., 2021. The Genetic Architecture of Obsessive-Compulsive Disorder: Contribution of Liability to OCD From Alleles Across the Frequency Spectrum. AJP appi.ajp.2021.21010101. https://doi.org/10.1176/appi.ajp.2021.21010101

Maj, M., Starace, F., Pirozzi, R., 1991. A family study of DSM-III-R schizoaffective disorder, depressive type, compared with schizophrenia and psychotic and nonpsychotic major depression. Am J Psychiatry 148, 612–616. https://doi.org/10.1176/ajp.148.5.612

Maxwell, M., 1996. Family Interview for genetic studies (FIGS): A manual for FIGS. Clinical Neurogenetics Branch, Intramural Research Program, National Institute for Mental Health, Bethesda, Maryland.

Meier, S.M., Petersen, L., Pedersen, M.G., Arendt, M.C.B., Nielsen, P.R., Mattheisen, M., Mors, O., Mortensen, P.B., 2014. Obsessive-compulsive disorder as a risk factor for schizophrenia: a nationwide study. JAMA Psychiatry 71, 1215–1221. https://doi.org/10.1001/jamapsychiatry.2014.1011

Narayanaswamy, J.C., Viswanath, B., Veshnal Cherian, A., Bada Math, S., Kandavel, T., Janardhan Reddy, Y.C., 2012. Impact of age of onset of illness on clinical phenotype in OCD. Psychiatry Res 200, 554–559. https://doi.org/10.1016/j.psychres.2012.03.037

Ozdemir, O., Tükel, R., Türksoy, N., Uçok, A., 2003. Clinical characteristics in obsessive-compulsive disorder with schizophrenia. Compr Psychiatry 44, 311–316. https://doi.org/10.1016/S0010-440X(03)00093-2

Pain, O., Hodgson, K., Trubetskoy, V., Ripke, S., Marshe, V.S., Adams, M.J., Byrne, E.M., Campos, A.I., Carrillo-Roa, T., Cattaneo, A., Als, T.D., Souery, D., …, Sullivan, P.F., McIntosh, A.M., Lewis, C.M., 2021. Identifying the Common Genetic Basis of Antidepressant Response. Biological Psychiatry: Global Open Science. https://doi.org/10.1016/j.bpsgos.2021.07.008

Pavuluri, M.N., Herbener, E.S., Sweeney, J.A., 2004. Psychotic symptoms in pediatric bipolar disorder. J Affect Disord 80, 19–28. https://doi.org/10.1016/S0165-0327(03)00053-3

Perris, F., Fabrazzo, M., De Santis, V., Luciano, M., Sampogna, G., Fiorillo, A., Catapano, F., 2019. Comorbidity of Obsessive-Compulsive Disorder and Schizotypal Personality Disorder: Clinical Response and Treatment Resistance to Pharmacotherapy in a 3-Year Follow-Up Naturalistic Study. Front Psychiatry 10, 386. https://doi.org/10.3389/fpsyt.2019.00386

Poyurovsky, M., Faragian, S., Pashinian, A., Heidrach, L., Fuchs, C., Weizman, R., Koran, L., 2008. Clinical characteristics of schizotypal-related obsessive-compulsive disorder. Psychiatry Res 159, 254–258. https://doi.org/10.1016/j.psychres.2007.02.019

Poyurovsky, M., Kriss, V., Weisman, G., Faragian, S., Schneidman, M., Fuchs, C., Weizman, A., Weizman, R., 2005. Familial aggregation of schizophrenia-spectrum disorders and obsessive-compulsive associated disorders in schizophrenia probands with and without OCD. Am J Med Genet B Neuropsychiatr Genet 133B, 31–36. https://doi.org/10.1002/ajmg.b.30148

Qin, H., Samuels, J.F., Wang, Y., Zhu, Y., Grados, M.A., Riddle, M.A., Greenberg, B.D., Knowles, J.A., Fyer, A.J., McCracken, J.T., Murphy, D.L., Rasmussen, S.A., Cullen, B.A., Piacentini, J., Geller, D., Stewart, S.E., Pauls, D., Bienvenu, O.J., Goes, F.S., Maher, B., Pulver, A.E., Valle, D., Lange, C., Mattheisen, M., McLaughlin, N.C., Liang, K.-Y., Nurmi, E.L., Askland, K.D., Nestadt, G., Shugart, Y.Y., 2016. Whole-genome association analysis of treatment response in obsessive-compulsive disorder. Mol. Psychiatry 21, 270–276. https://doi.org/10.1038/mp.2015.32

Rajkumar, R.P., Reddy, Y.C.J., Kandavel, T., 2008. Clinical profile of “schizo-obsessive” disorder: a comparative study. Compr Psychiatry 49, 262–268. https://doi.org/10.1016/j.comppsych.2007.09.006

Sheehan, D.V., Lecrubier, Y., Sheehan, K.H., Amorim, P., Janavs, J., Weiller, E., Hergueta, T., Baker, R., Dunbar, G.C., 1998. The Mini-International Neuropsychiatric Interview (M.I.N.I.): the development and validation of a structured diagnostic psychiatric interview for DSM-IV and ICD-10. J Clin Psychiatry 59 Suppl 20, 22-33;quiz 34-57.

Smoller, J.W., Andreassen, O.A., Edenberg, H.J., Faraone, S.V., Glatt, S.J., Kendler, K.S., 2019. Psychiatric genetics and the structure of psychopathology. Mol Psychiatry 24, 409–420. https://doi.org/10.1038/s41380-017-0010-4

Sreeraj, V.S., Holla, B., Ithal, D., Nadella, R.K., Mahadevan, J., Balachander, S., Ali, F., Sheth, S., Narayanaswamy, J.C., Venkatasubramanian, G., John, J.P., Varghese, M., Benegal, V., Jain, S., Reddy, Y.J., ADBS Consortium, Viswanath, B., 2020. Psychiatric symptoms and syndromes transcending diagnostic boundaries in multiplex families (preprint). medArXiv. https://doi.org/10.1101/2020.01.06.20016543

Swets, M., Dekker, J., van Emmerik-van Oortmerssen, K., Smid, G.E., Smit, F., de Haan, L., Schoevers, R.A., 2014. The obsessive compulsive spectrum in schizophrenia, a meta-analysis and meta-regression exploring prevalence rates. Schizophr Res 152, 458–468. https://doi.org/10.1016/j.schres.2013.10.033

Van Dael, F., van Os, J., de Graaf, R., ten Have, M., Krabbendam, L., Myin-Germeys, I., 2011. Can obsessions drive you mad? Longitudinal evidence that obsessive-compulsive symptoms worsen the outcome of early psychotic experiences. Acta Psychiatr Scand 123, 136–146. https://doi.org/10.1111/j.1600-0447.2010.01609.x

Vandeleur, C.L., Merikangas, K.R., Strippoli, M.-P.F., Castelao, E., Preisig, M., 2014. Specificity of psychosis, mania and major depression in a contemporary family study. Mol Psychiatry 19, 209–213. https://doi.org/10.1038/mp.2013.132

Viswanath, B., Narayanaswamy, J.C., Rajkumar, R.P., Cherian, A.V., Kandavel, T., Math, S.B., Reddy, Y.C.J., 2012. Impact of depressive and anxiety disorder comorbidity on the clinical expression of obsessive-compulsive disorder. Compr Psychiatry 53, 775–782. https://doi.org/10.1016/j.comppsych.2011.10.008

Viswanath, B., Rao, N.P., Narayanaswamy, J.C., Sivakumar, P.T., Kandasamy, A., Kesavan, M., Mehta, U.M., Venkatasubramanian, G., John, J.P., Mukherjee, O., Purushottam, M., Kannan, R., Mehta, B., Kandavel, T., Binukumar, B., Saini, J., Jayarajan, D., Shyamsundar, A., Moirangthem, S., Vijay Kumar, K.G., Thirthalli, J., Chandra, P.S., Gangadhar, B.N., Murthy, P., Panicker, M.M., Bhalla, U.S., Chattarji, S., Benegal, V., Varghese, M., Reddy, J.Y.C., Raghu, P., Rao, M., Jain, S., 2018. Discovery biology of neuropsychiatric syndromes (DBNS): a center for integrating clinical medicine and basic science. BMC Psychiatry 18, 106. https://doi.org/10.1186/s12888-018-1674-2

Zhang, R., Kuja-Halkola, R., Birgegård, A., Larsson, H., Lichtenstein, P., Bulik, C. M., & Bergen, S. E. 2021. Association of family history of schizophrenia and clinical outcomes in individuals with eating disorders. Psychological Med, 1–8. Advance online publication. https://doi.org/10.1017/S0033291721001574

